# Autoimmune anti-DNA antibodies predict disease severity in COVID-19 patients

**DOI:** 10.1101/2021.01.04.20249054

**Authors:** Claudia Gomes, Marisol Zuniga, Kelly A. Crotty, Kun Qian, Lawrence Hsu Lin, Kimon V. Argyropoulos, Huilin Li, Paolo Cotzia, Ana Rodriguez

**Author notes:** Both authors contributed equally (Claudia Gomes, Marisol Zuniga).

## Abstract

COVID-19 can lead to severe disease and death, however the mechanisms of pathogenesis in these patients remain poorly understood. High levels of autoimmune antibodies have been observed frequently in COVID-19 patients but their specific contribution to disease severity and clinical manifestations remain unknown.

We performed a retrospective study of 115 COVID-19 hospitalized patients with different degrees of severity to analyze the generation of autoimmune antibodies to common antigens: a lysate of erythrocytes, the lipid phosphatidylserine (PS) and DNA.

High levels of IgG autoantibodies against erythrocyte lysates were observed in a large percentage (up to 41%) of patients. Anti-DNA antibodies determined upon hospital admission correlated strongly with later development of severe disease, showing a positive predictive value of 89.5% and accounting for 22% of total severe cases. Statistical analysis identified strong correlations between anti-DNA antibodies and markers of cell injury, coagulation, neutrophil levels and erythrocyte size.

Anti-DNA autoantibodies may play an important role in the pathogenesis of COVID-19 and could be developed as a predictive biomarker for disease severity and specific clinical manifestations.

## Introduction

Infections trigger immune responses that target pathogen antigens, but frequently they also induce potent autoimmune responses that are characterized by high levels of antibodies recognizing a variety of host antigens (1). Autoimmune antibodies have been characterized in viral diseases such as hepatitis C, HIV, and arboviral infections, but also in bacterial and protozoan infections like tuberculosis, and malaria. Infection-induced autoantibodies can recognize a variety of self-antigens, including nucleic acids, lipids, and glycoproteins (1).

Autoimmune antibodies can contribute to systemic inflammatory responses and subsequent tissue damage through different mechanisms, including immune-complex formation, complement activation, formation of thrombi and/or lysis of uninfected cells (2).

In COVID-19, autoantibodies are found in high levels in a large proportion of hospitalized patients with severe disease (3-5) and have been associated with the development of autoimmune pathologies (6), such as thrombocytopenia (7), hemolytic anemia (8), Guillain-Barre (9), and anti-phospholipid (10, 11) syndromes. Autoantibodies against type I interferons that neutralize the anti-viral activity of these cytokines have also been identified and may explain disease severity among a subset of COVID-19 patients (12).

In addition to acute respiratory distress and pulmonary edema, COVID-19 can cause multi-organ widespread thrombosis and disseminated intravascular coagulation (13). To explore the relation of autoimmune antibodies to COVID-19 clinical manifestations, we have determined IgG autoantibody levels in COVID-19 patients’ plasma against a lysate of erythrocytes (as a general measure of autoreactivity) and two specific antigens that are involved in the autoimmune pathogenesis of other diseases: anti-phosphatidylserine (PS) (14) and anti-DNA (15). Our analysis showed that some hospitalized patients present high levels of these autoantibodies and that there is a strong correlation among these levels in patients, pointing to general autoreactivity as a common phenomenon in COVID-19 patients. Among hospitalized patients, severity of disease was strongly correlated with levels of anti-DNA antibodies and weakly with anti-PS. High-throughput data analysis showed that anti-DNA antibodies correlate strongly with parameters related to cell injury, coagulation, neutrophil levels and erythrocyte distribution width, suggesting a role of these autoantibodies in exacerbating COVID-19 clinical course of disease.

## Methods

### Bioethics statement

The collection of COVID-19 human biospecimens for research has been approved by NYULH Institutional Review Board under the S16–00122 Universal Mechanism of human bio-specimen collection and storage for research. This protocol allows the collection and analysis of clinical and demographic data. The database used for this project is de-identified.

### Study design and participants

This retrospective cohort study included 115 hospitalized cases of SARS-CoV-2 infection at NYU Langone Health (NYULH) and 20 uninfected controls. All COVID-19 cases occurred from April until June 2020, during the peak of the COVID-19 pandemic in this area. Control samples used were collected earlier than January 2020, to avoid possible undiagnosed COVID-19 cases. Patients that needed intubation or required intensive care monitoring were considered severe.

### Sample collection and testing for COVID-19

Sample collection was performed as part of routine clinical care for COVID-19 patients at NYULH at the time of hospital admission (day 0 or 1). Venous blood samples were collected in standard plasma separation tubes containing heparin (16 USP units/ml) (BD Diagnostics). Plasma was recovered following centrifugation, stored at 5°C for 5 days (in case further clinical testing for the patient was required) before aliquoting and storage at −80°C. Assessment of SARS-CoV-2 infection was performed using nasopharyngeal swabs in clinical PCR assays as described (16).

### Clinical parameters analysis

Every patient sample is associated with a list of parameters, including non-identifiable information (age, sex, race), clinical data and complete immunological, hematological, and biochemical clinical characterization. The clinical parameters used for the analysis had data from more than 30 patients (n > 30). The analysis was performed to determine statistical correlations between the levels of autoantibodies, that were determined on day 0-1 of admission and clinical parameters determined on day 0-3, to include some tests that were performed a few days after admission. A parallel analysis was also performed to analyze statistical significance in the relation between autoantibody levels and the minimum or maximum value obtained for every laboratory test during each patient stay at the hospital, with the aim to determine the predictive value of autoantibody levels for the different clinical parameters.

### Data analysis

SQL statements were generated programmatically to select specific records from NYU deidentified COVID-19 database. WHERE clauses were generated so that only specific patient records were selected. Records of interest were then exported to Excel using the HUE SQL query tool. Excel worksheets were generated using the Infragistics Excel Engine™ software library (Infragistics, Inc., Cranbury, NJ) which synchronized the data records obtained from the NYU COVID-19 database with deidentified ID numbers which correspond to patient samples. A simple algorithm was applied to the lab result records to obtain the minimum and maximum values for each lab test and create new worksheets, designed to be consumed by statistical analysis software.

### Statistical analysis

Data were analyzed using GraphPad Prism v8. Two tailed Spearman correlation was used to evaluate the association between the different autoantibodies and between the autoantibodies and clinical parameters. False discovery rate (FDR) was controlled for multiple comparisons within specific groups. The differences in autoantibody levels between controls and COVID-19 patients were determined using Mann-Whitney test. The association between living status and severity of disease with the autoantibodies was performed using logistic regression, adjusting for age, race, and gender.

### ELISA

To measure autoimmune antibodies, Immulon 2HB 96-well ELISA plates were coated with lysates of human erythrocytes (2×10^3^ cells/µl in PBS, from Interstate Blood Bank), PS (Sigma) at 20 μg/ml in 200-proof Molecular Biology ethanol, calf thymus DNA (predominantly double stranded, Sigma) at 10 μg/ml in PBS. Plates were allowed to evaporate for 16 h at 4°C. PS-coated plates were let evaporate at RT until completely dry. Plates were washed 3 times with PBS 0.05% Tween-20 and then blocked for 1 h at 37°C with PBS 3% BSA. Plasma from patients or control was diluted at 1:100 in blocking buffer and incubated in duplicates for 2 h at 37°C. Plates were washed again 3 times and incubated with a polyclonal goat anti-human IgG-HRP diluted 1:500 (Invitrogen) for 1 h at 37°C. Plates were washed 4 more times and developed using TMB substrate (BD Biosciences). The reaction was stopped using Stop buffer (Biolegend) and absorbance read at 450 nm. The mean OD at 450 nm from duplicate wells was compared with a reference positive control plasma sample, previously identified as high responder for IgG autoantibodies, to calculate relative units (RU). To determine IC, plates were coated with C1q polyclonal antibody (Invitrogen, 1μg/ml in carbonate-bicarbonate buffer) and the same protocol was followed with the following modifications: plates were blocked for 1.5 h with PBS 0.1% BSA, plasma and secondary antibody were diluted 1:80 and 1:5000, respectively. All steps were performed at room temperature. Samples were considered positive for autoantibodies if the relative units (RU) were greater than the mean plus 3 times the standard deviation of the controls.

### Role of the funding source

The funder had no role in study design, data collection, data analysis, data interpretation, or writing of the report. The corresponding author had full access to all data in the study and had final responsibility for the decision to submit for publication.

## Results

### Patients characteristics

115 cases of hospitalized patients with SARS-CoV-2 infection with different degrees of disease severity and 20 uninfected controls were used in this retrospective study. Patients were stratified into three different groups: non-severe, severe patients that survived and severe patients that died of COVID-19 (Table I).

**Table I.** Demographic and clinical characteristics of patients (n = 115).

|  |  |
| --- | --- |
| Age years, median (IQR) | 66 (20-101) |
| Female sex, n (%) | 35 (30.4) |
| Race, n (%) |  |
| White | 68 (59.1) |
| Black | 14 (12.2) |
| Asian | 8 (7.0) |
| Other | 25 (21.7) |
| Disease severity, n (%) |  |
| Non-severe | 40 (34.8) |
| Severe that survived | 42 (36.5) |
| Severe that died | 33 (28.7) |

### Autoantibody levels are elevated in hospitalized patients with COVID-19 compared to controls

The plasma of controls and COVID-19 patients on day 0-1 was used to determine the levels of different IgG autoimmune antibodies. As a general measurement of the autoimmune antibody response, samples were tested for reactivity to a lysate of human erythrocytes (RBCL). Autoantibodies to the phospholipid PS and DNA were also determined, since they have been involved in the pathogenesis of other diseases (14, 15).

We observed that COVID-19 patients had significantly higher average levels of circulating anti-RBCL and anti-PS compared to controls, with 41% of patients testing positive for anti-RBCL (Fig. 1A). The levels of autoantibodies increased with severity of disease, showing higher levels in severe patients compared to non-severe or control (Fig. 1B).

**Figure 1.**
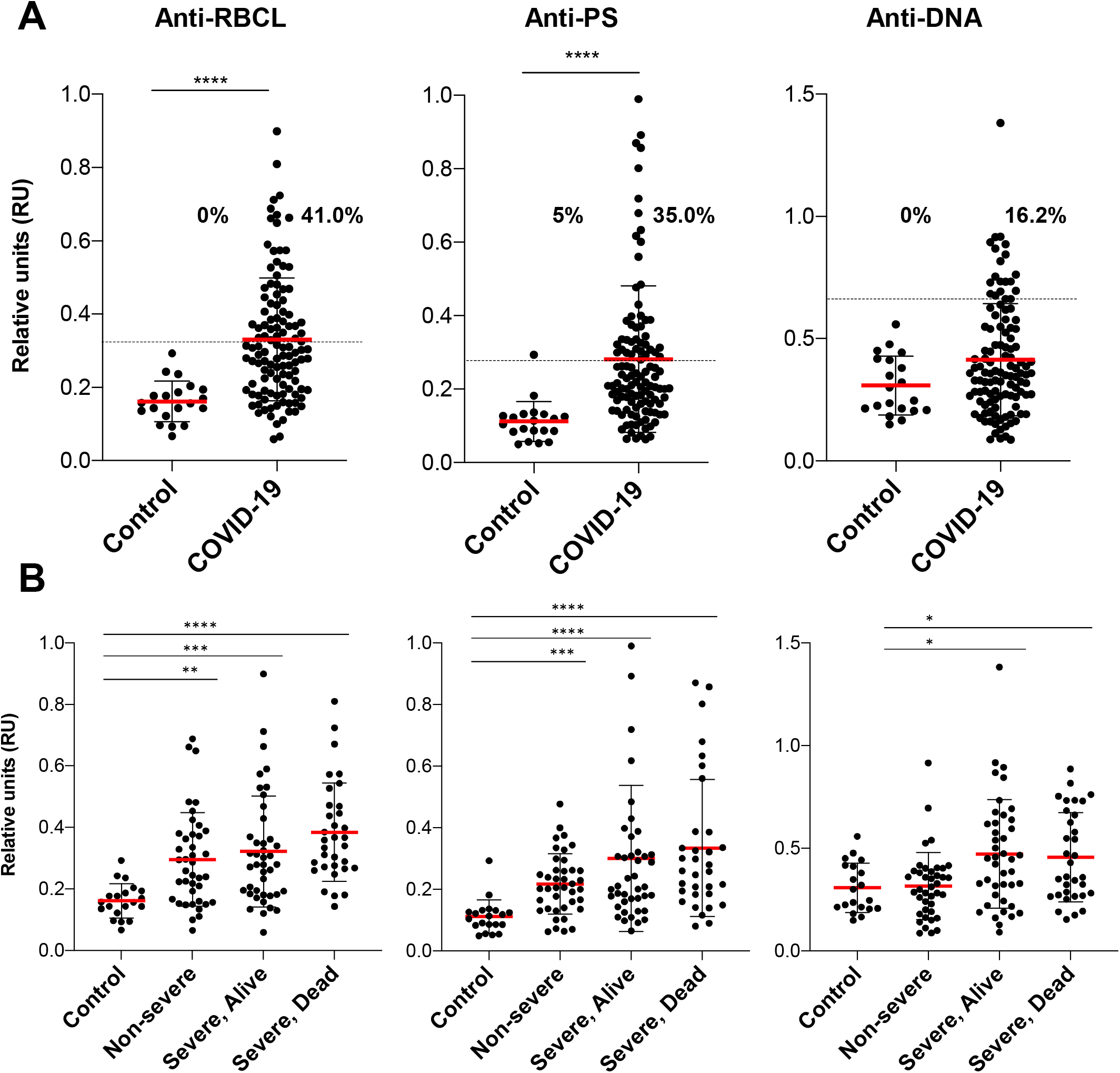
COVID-19 patients present higher levels of IgG autoimmune antibodies compared to uninfected controls. (A) Analysis of 115 plasma samples of COVID-19 patients and 20 controls for levels of IgG, anti-RBCL, anti-PS and anti-DNA. Samples were considered positive for autoantibodies if the relative units (RU) > the mean plus 3 times the standard deviation of the controls. Percentage of positive samples for each group is indicated. The cut-off is indicated by the dashed horizontal line. (B) Comparison of controls and COVID-19 patients stratified by severity of disease. Average of duplicated values for each patient are shown. *p<0.05, **p<0.001, ***p<0.0001**** p<0.0001, Mann-Whitney test.

In patients with autoimmune diseases, binding of autoantibodies to their antigens results in the formation of immune-complexes (IC), which are deposited in various tissues frequently leading to disease (2). The levels of circulating IC were determined in this cohort, finding no significant differences between controls and COVID-19 patients (Fig. 2).

**Figure 2.**
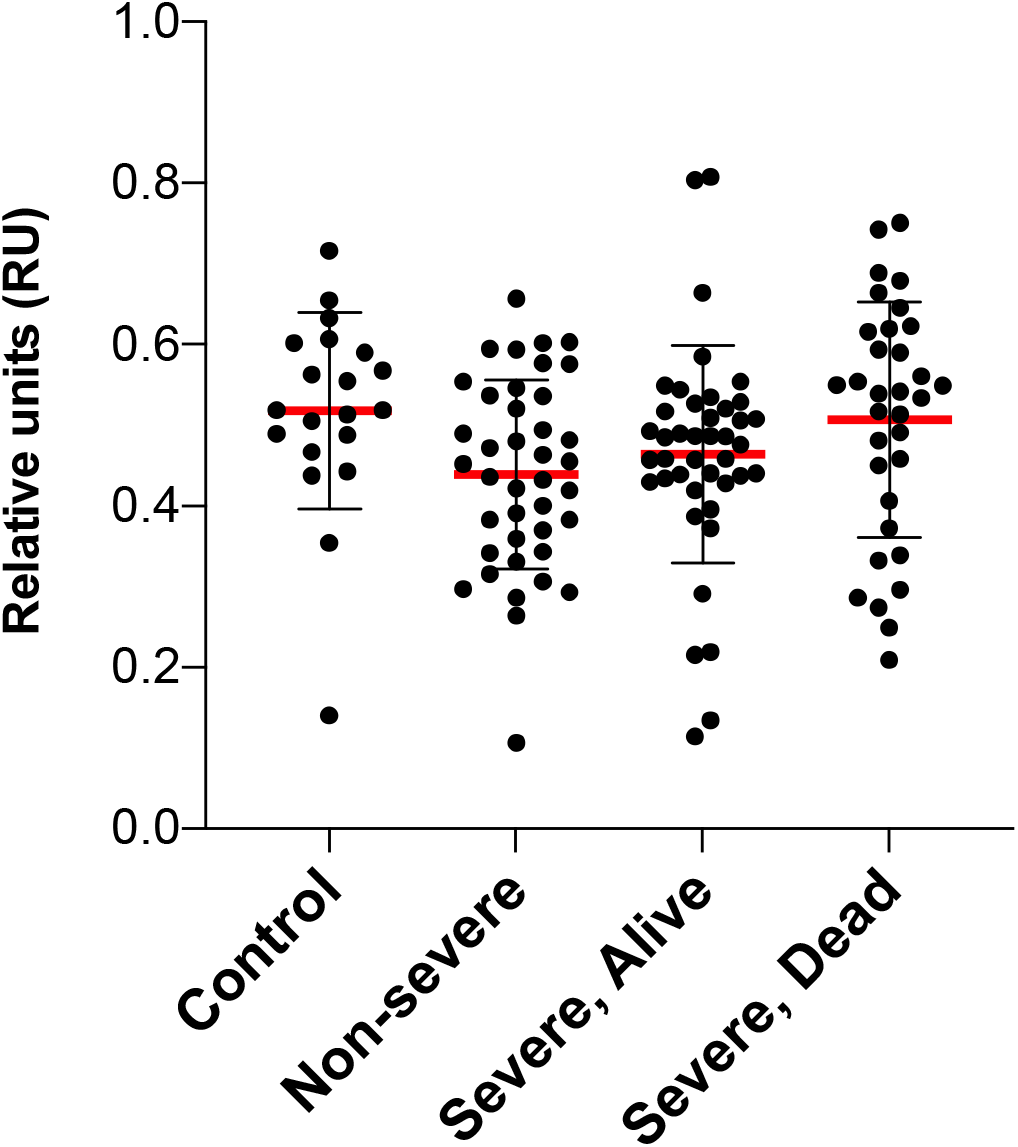
COVID-19 patients do not present higher levels of IC. Analysis of 115 plasma samples of COVID-19 patients stratified by severity of disease and 20 controls for levels of C1q-binding IC.

The levels of the three different autoantibodies that we examined are highly correlated with each other (Table II), indicating that individual patients tend to have similar levels of different autoantibodies and suggesting some patients are more prone to autoreactivity.

**Table II.**
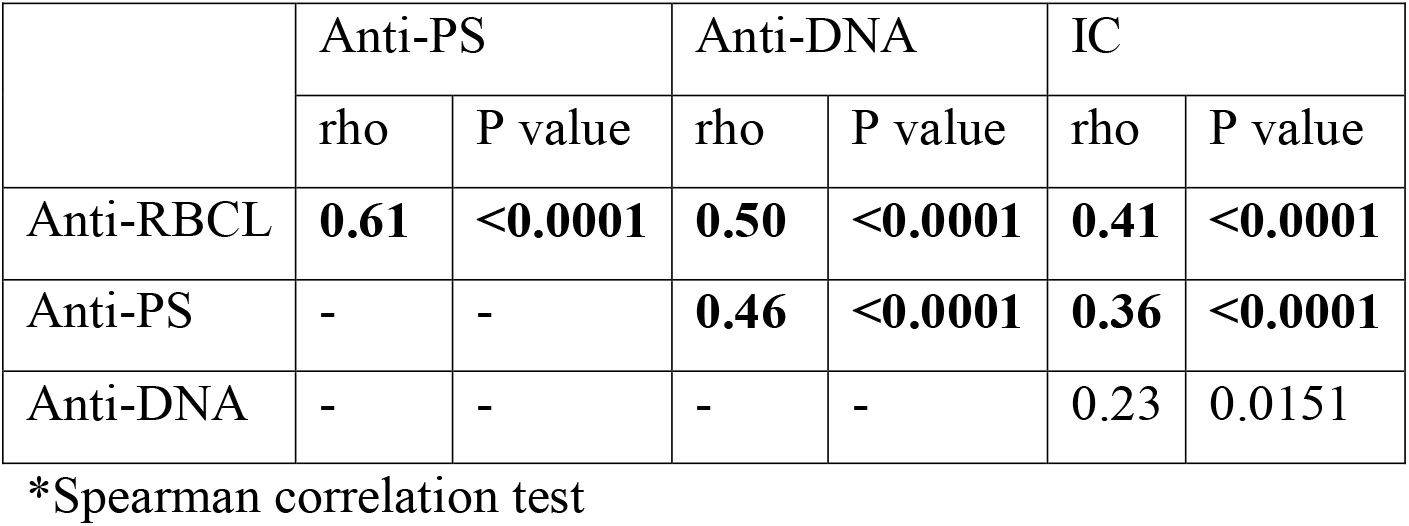
Correlation among autoantibody and immune-complex levels.

|  | Anti-PS |  | Anti-DNA |  | IC |  |
| --- | --- | --- | --- | --- | --- | --- |
|  | rho | P value | rho | P value | rho | P value |
| Anti-RBCL | **0.61** | **<0.0001** | **0.50** | **<0.0001** | **0.41** | **<0.0001** |
| Anti-PS | - | - | **0.46** | **<0.0001** | **0.36** | **<0.0001** |
| Anti-DNA | - | - | - | - | 0.23 | 0.0151 |

\*Spearman correlation test

When the severity of disease or death occurrence were analyzed in regard to the levels of autoantibodies, we observed a strong correlation of anti-DNA antibody levels with disease severity (OR = 7.2, p = 0.006, after adjustment for age, race, and sex) (Table III).

**Table III.** Correlation analysis between autoantibodies and severity/living status of hospitalized patients.

|  | **Odds**<br>**ratio\*\*** | **p**<br>**value\*\*\*** |
| --- | --- | --- |
| Living status* |  |  |
| anti-RBCL | 3.921 | 0.137 |
| anti-PS | 2.083 | 0.161 |
| anti-DNA | 1.925 | 0.161 |
| IC | 4.759 | 0.137 |
| Severity* |  |  |
| anti-RBCL | 2.741 | 0.119 |
| anti-PS | 5.765 | 0.043 |
| **anti-DNA** | **7.219** | **0.006** |
| IC | 3.477 | 0.119 |

\*Regression analysis adjusted for age, sex and race.
\*\*1 unit = 0.5 RU
\*\*\*FDR adjusted p

We observed that a large proportion of patients that were positive for anti-DNA antibodies upon hospital admission developed severe disease. Anti-DNA antibodies showed a positive predictive value of 89% for COVID-19 severity (Table IV), which corresponds to 22% of total severe cases in this cohort (17 out of 75).

**Table IV.** Autoantibodies positive predictive value (PPV) for disease severity in hospitalized patients.

|  | PPV, %<br>(positives/total)* |
| --- | --- |
| anti-RBCL | 64.6 (31/48) |
| anti-PS | 78.0 (32/41) |
| anti-DNA | 89.5 (17/19) |
| IC | 0 (0/0) |

\*Patients that develop severe disease from total autoantibody or IC positive at day 0

### Relationship between autoantibody levels and clinical parameters

We first performed a statistical analysis comparing the levels of the three determined autoimmune IgG antibodies and IC with values of laboratory tests assessed in the 115 patients hospitalized in the cohort. These include data from 118 parameters that were assessed in the context of hematologic, metabolic, immunological, and biochemical monitoring of these patients during their admission.

No significant correlations were found between the levels of any of the autoantibodies or IC with any of the clinical parameters measured at day 0-3 (Table S1). However, significant correlations (rho > 0.3 and p < 0.01) were found between the levels of different autoantibodies and the maximum value of several laboratory tests performed during each patient stay at the hospital. The correlations of each autoantibody type were distinct from the others and tended to be clustered in groups of parameters related to specific disease processes (Fig. 3, Table S2). Analysis of the minimum values of laboratory tests did not identify any significant correlations with autoantibody levels (Table S3).

**Figure 3.**
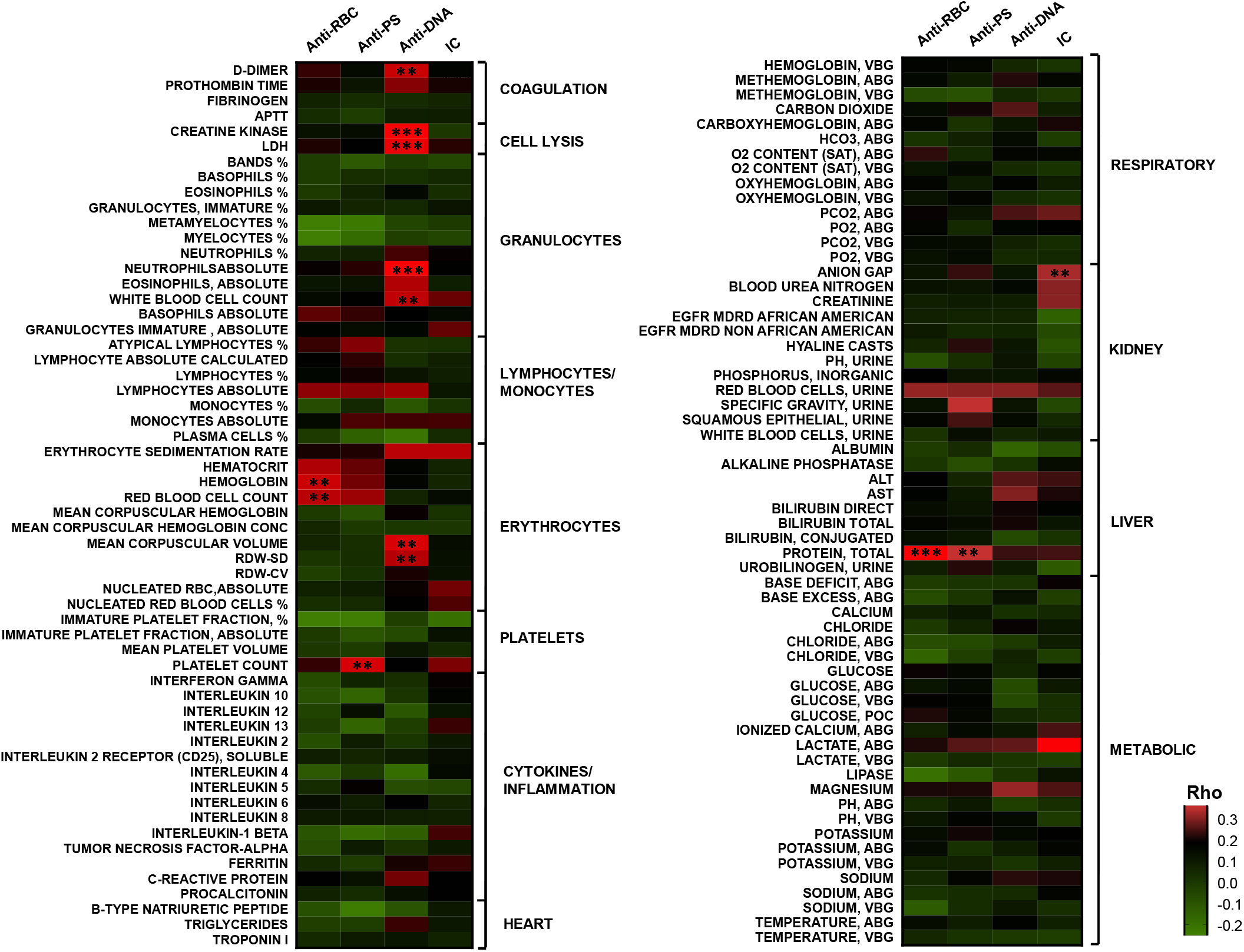
Correlation analysis of autoantibodies and maximum values of clinical tests identified significant associations mainly for anti-DNA antibodies. Heatmap listing 118 clinical tests correlation with the autoantibodies and IC. Color scale indicates rho values for each pair in the two tailed Spearman correlation test. Corresponding p-value (with FDR correction) is indicated if p < 0.001 (**p<0.001, ***p<0.0001).

Anti-DNA antibodies correlated strongly with the maximum values of two markers of cellular injury: lactate dehydrogenase (LDH), which is released by all cell types, and creatine kinase, which is released specifically by striated muscle cells, suggesting a possible link between anti-DNA antibodies and cellular lysis (Fig. 3). Significant correlations were also found between anti-DNA antibodies and absolute numbers of neutrophils, total white blood cell counts and markers of RBCs volume, such as mean corpuscular volume and red cell distribution width (RDW). We also observed that maximum values of D-dimer concentration, a biomarker for coagulation disorders and thrombus formation, correlated significantly with the levels of anti-DNA antibodies.

Anti-RBCL and anti-PS antibodies correlated only with the total levels of protein in plasma and parameters related to RBC and platelet levels, respectively. IC levels correlated with parameters related to kidney function, as it is typical in autoimmune disorders.

It is important to note that no correlation was found between any of the autoantibodies and cytokines or inflammatory mediators such as C-reactive protein, several interleukins (including IL6), TNF or interferon-γ.

## Discussion

Although autoantibodies have been observed in different viral and non-viral infections (1), the high percentage of COVID-19 patients with autoantibodies (up to 33% positive for anti-RBCL) and the strong correlation of anti-RBCL autoantibodies and total protein levels in plasma, indicates that the levels of autoreactivity are particularly high in this infection. Similarly, a study of autoantibodies to protein antigens in COVID-19 patients also found high general autoreactivity (3).

We observed that the levels of the three autoantibodies determined are highly correlated with each other, which is consistent with the activation of a generalized autoimmune response in certain patients during COVID-19. However, we observed a higher proportion of patients with elevated levels of circulating anti-RBCL and anti-PS compared to anti-DNA, which may suggest that specific autoantibodies are produced more frequently in our cohort, but also that antibodies with certain specificities may be removed from the circulation more efficiently.

In contrast to other infectious diseases that also induce autoantibodies (1), we observed that the levels of IC are not increased in the circulation of COVID-19 patients. This is unexpected, considering the high levels of autoimmune antibodies with specificities for antigens that are likely to be accessible in the circulation, such as PS in exosomes or cell-free DNA (17). However, it is possible that IC are being formed, but are rapidly removed from the circulation, which would be consistent with the observed correlations between levels of IC and parameters indicating kidney disfunction.

Other studies have analyzed the relation of autoantibodies of different specificities and disease severity, finding that protein antigens such as type I interferons (12) or particular tissue associated antigens (3) correlate with COVID-19 severity in specific subgroups of patients.

Our study is focused on two specific autoantibodies, anti-DNA and anti-PS, because of their previously described roles in the pathogenesis of other diseases (14, 15) and their capacity to bind to different cell types in the circulation (18, 19). We observed that anti-DNA antibodies, which are typically found in autoimmune disorders, such as Systemic Lupus Erythematosus (20) and in some infectious diseases (21), are found in 13% of COVID-19 patients in this cohort and correlate strongly with severity of COVID-19 disease in this group. Since the levels of anti-DNA were determined upon hospital admission, it is possible that this simple test could be developed as a predictor of COVID-19 severity. In this cohort, high anti-DNA antibodies accounted for 22% of total severe patients.

COVID-19 patients have high levels of cell-free DNA in the circulation, which are also related to disease severity (17, 22). Furthermore, we observed a strong correlation between markers of cell injury and anti-DNA levels, which mirrors the previously found correlation between these markers and cell-free DNA levels (17). Since the levels of cell-free DNA and anti-DNA correlate with severity of disease and markers of cell injury, it is likely that the binding of anti-DNA to cell-free DNA contributes to the pathogenesis of COVID-19 manifestations.

It is known that cell-free DNA binds to the surface of endothelial and immune cells (19), as well as erythrocytes (23), where it could constitute a target for anti-DNA antibodies in the circulation, triggering complement-mediated cell lysis. We observed a strong correlation of anti-DNA antibodies with lactate dehydrogenase (LDH) and creatine kinase, suggesting that anti-DNA antibodies may contribute to muscle injury, which is frequent in COVID-19 patients (24).

High levels of cell-free DNA in COVID-19 patients are probably a result of the active release of Neutrophil Extracellular Traps (NETs) which are mainly composed of neutrophil DNA (25). Interestingly, anti-DNA antibodies also correlate with absolute levels of neutrophils.

Anti-DNA antibodies also correlate with a marker of thrombosis, D-dimer, but not with other parameters involved in triggering the coagulation cascade. Since NETs contribute to thrombosis and have been found to be components of the micro-thrombi in COVID-19 patients (25), it is possible that anti-DNA antibodies bind to the DNA in NETs, facilitating cellular aggregation and possibly contributing to intravascular coagulation.

Anti-DNA antibodies also correlate with a parameter that determines the variation of erythrocyte volume, RDW, which has been associated with mortality risk in multiple diseases, including COVID-19 (26).

Severe complications in COVID-19 patients typically develop at least one week after the onset of symptoms (27), when viral levels are already decreasing and are frequently undetectable (28). These observations suggest that the most severe forms of disease observed in COVID-19 patients may be a result of the host response to infection, rather than a direct consequence of viral cytopathic effect. Autoantibodies, as part of the host response to infection, may contribute to this delayed pathogenesis through different mechanisms. Our work suggests that anti-DNA antibodies may have a role in different pathogenic processes, including cell injury and coagulation, constituting a possible mechanism contributing to pathogenesis in COVID-19 patients.

## Data Availability

All data referred to in the manuscript is available in the provided supplementary tables

## Contributors

MZ, CG and KAC performed all the experimental determinations and processed the data. KQ and HL performed the statistical analysis of the data. LHL, KVA, PC helped with the collection of samples, performed case identification and revised the manuscript. AR had the idea for the study and contributed to data analysis, writing, and editing the manuscript.

## Declaration of interests

The authors declare that they have no conflict of interest.

## Acknowledgements

We thank the personnel at the NYU School of Medicine Center for Biospecimen Research and Development (CBRD) and Brian Fallon for bioinformatics support.

## Funding

This research was supported by the NYU Langone COVID-19 special fund.

## Notes

### Competing Interest Statement

The authors have declared no competing interest.

### Funding Statement

This research was supported by the NYU Langone COVID-19 special fund. The funder had no role in study design, data collection, data analysis, data interpretation, or writing of the report.

### Author Declarations

The collection of COVID-19 human biospecimens for research has been approved by NYULH Institutional Review Board under the S16-00122 Universal Mechanism of human bio-specimen collection and storage for research. This protocol allows the collection and analysis of clinical and demographic data. The database used for this project is de-identified.

